# Ready for a BASE jump? Do not neglect SARS-CoV-2 hospitalization and fatality risks in the middle-aged adult population

**DOI:** 10.1101/2020.11.06.20227025

**Authors:** Nathanael Lapidus, Juliette Paireau, Daniel Levy-Bruhl, Xavier de Lamballerie, Gianluca Severi, Mathilde Touvier, Marie Zins, Simon Cauchemez, Fabrice Carrat, the SAPRIS-SERO study group

## Abstract

Seroprevalence results coupled with surveillance data were used to estimate the SARS-CoV-2 infection hospitalization (IHR) and infection fatality ratios (IFR) in France. IHR and IFR were dramatically high in the very elderly (80-90 years: IHR: 30%, IFR: 11%), but also substantial in middle-aged adults (40-50 years: IHR: 1.2%, IFR: 0.05%).

## Introduction

The SARS-CoV-2 infection hospitalization ratio (IHR, probability of hospitalization in infected individuals) and the infection fatality ratio (IFR, probability of death in infected individuals) are critical parameters in public health decision-making regarding prioritization of control measures. However, age- and sex-related estimates are scarce, as they need reliable cumulative estimates of past infections, hospitalizations and deaths. By May 11, 2020, the cumulative number of COVID-19-related hospitalizations and hospital deaths in France reached 96,000 and 17,000, respectively. This study estimated the age- and sex-specific IFR and IHR in France for this period based on contemporary SARS-CoV-2 seroprevalence data.

## Methods

Data from a seroprevalence study performed in May-June 2020 in 20-to 90-year old subjects were used. This study included subjects from three pre-existing general adult population cohorts from Île-de-France (N=6,348) and Grand Est (N=3,434), two regions of France with the highest incidence of COVID-19 during the first wave of the pandemic. (1) All participants from these cohorts with regular access to online questionnaires were invited to participate in the study and dried-blood spots were collected in a random sample among them. All samples were processed with an ELISA test to detect antibodies directed against the spike protein, and for some of them with an ELISA test to detect antibodies directed against the nucleocapsid protein and a micro-neutralization assay to detect neutralizing antibodies (technical details were reported elsewhere (1)). Serological status was estimated with a multiple imputation approach relying on participants’ age, sex and serological results for all available tests. Seroprevalence estimates were calibrated by generalized raking in relation to census data from the general adult population, excluding nursing home residents who were not part of the cohort target population. The cumulative numbers of hospital admissions and deaths for COVID-19 were obtained from the SI-VIC database, the national exhaustive inpatient surveillance system used during the pandemic. Patients from nursing homes were removed from these counts. Since the median date of sample collection in the serosurvey was May 14, hospital admissions and deaths were considered up to May 6 and May 13 to account for estimated 11- and 19-day delays from infection to hospitalization and seroconversion, respectively, (2,3) and an estimated 7-day delay from hospitalization to death (SI-VIC data (4)). We report IHR and IFR by sex and 10-year age class. Multivariable random-effect meta-regression models were fitted to estimate the relative risks (RR) of hospitalization and death according to age and sex, using age class as a continuous covariate.

## Results

The overall estimated IHR and IFR in the French adult population (excluding nursing homes) were 3.2% (95% confidence interval (CI): 2.8, 3.6) and 0.58% (95% CI: 0.52, 0.65), respectively. We found a strong log-linear relationship between age and the risk of hospitalization or death, corresponding to an exponential increase in risk with age (Figure). The estimated IHR in 20-30, 40-50, 60-70 and 80-90-year-old subjects was 0.46% (95% CI: 0.30, 0.72), 1.2% (95% CI: 0.96, 1.4), 6.9% (95% CI: 5.3, 9.0) and 30% (95% CI: 10, 99), respectively (RR: 2.1 (95% CI: 1.9, 2.3) per 10-year increase). The estimated IFR in these same age groups was 0.0077% (95% CI: 0.0050, 0.012), 0.050% (95% CI: 0.041, 0.060), 1.0 (95% CI: 0.81, 1.4) and 11% (95% CI: 3.6, 35), respectively (RR: 3.8 (95% CI: 2.4, 4.2) per 10-year increase). Both IHR and IFR were higher in men than in women for all age classes: RR: 1.5 (95% CI: 1.1, 2.1) and 2.5 (95% CI: 1.8, 3.5), respectively.

**Figure 1:**
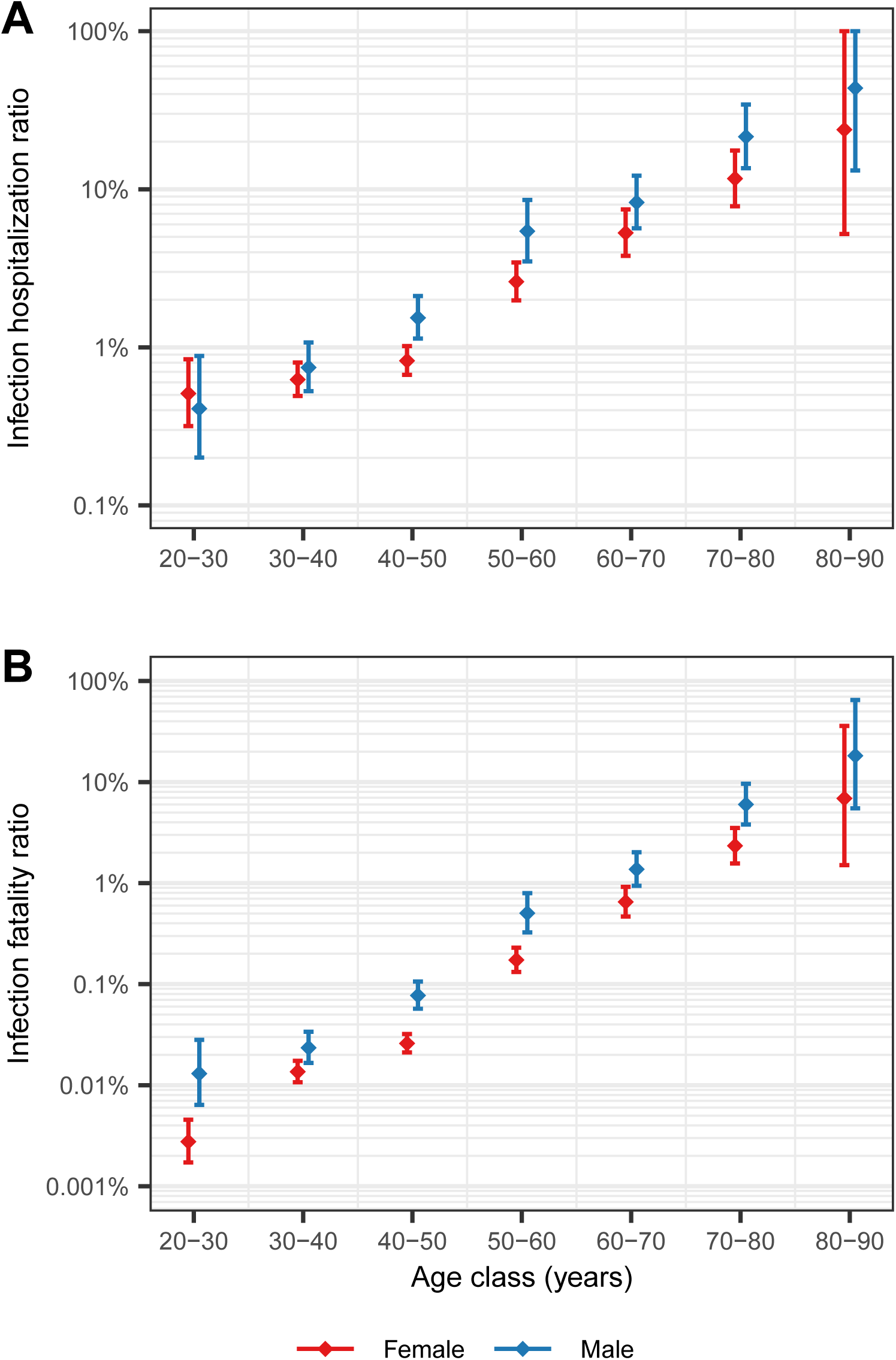
Infection hospitalization ratio and infection fatality ratio with 95% confidence intervals, by age class and sex, estimated from seroprevalence data in France (logarithmic scale). Panel A: Infection hospitalization ratio. Panel B: Infection fatality ratio

## Discussion

We identified an overall IHR of 3.2% and an IFR of 0.58% in the adult population, excluding those in nursing homes. Though these ratios may be underestimated in the elderly as nursing home residents are likely to experience poorer outcomes, our estimates are in line with earlier models (3) and various age-specific IFR estimates, worldwide. (5) The IFR exponentially increases with age (doubling every 5.2 years) and is higher in men. Both IHR and IFR were dramatically elevated in the very elderly but this should not obscure substantial estimates in the young or middle-aged adult population. For example, in 20-to 30-year-old adults, the risk of death is 8 times that of a skydiving jump (1 per 100,000 jumps (6)) and in 40 to 50-year-olds, this risk of death is close to that of a BASE jump (1 to 2 per 1000 jumps (6)). From a public health perspective, these findings support the need for comprehensive preventive measures to help reduce the spread of the virus, even in young or middle-aged adults.

## Supporting information

The SAPRIS-SERO study group

## Data Availability

Data are not publicly available. Access to the SI-VIC database must be granted by Sante Publique France and access to serosurvey data must be granted by the SAPRIS-SERO study group.

## Ethics approval

The SAPRIS-SERO study was approved by the Sud-Mediterranée III ethics committee (approval #20.04.22.74247) and electronic informed consent was obtained from all participants for DBS testing.

