## Supplementary material for "Ready for a BASE jump? Do not neglect SARS-CoV-2 hospitalization and fatality risks in the middle-aged adult population": The SAPRIS-SERO study group

Sofiane Kab, Adeline Renuy, Stephane Le-Got, Celine Ribet, Emmanuel Wiernik, Marcel Goldberg, Marie Zins (Constances cohort),

Fanny Artaud, Pascale Gerbouin-Rérolle, Mélody Enguix, Camille Laplanche, Roselyn Gomes-Rima, Lyan Hoang, Emmanuelle Correia, Alpha Amadou Barry, Nadège Senina, Gianluca Severi (E3N-E4N cohort)

Fabien Szabo de Edelenyi, Nathalie Druesne-Pecollo, Younes Esseddik, Serge Hercberg, Mathilde Touvier (NutriNet-Santé cohort)

Marie-Aline Charles, Pierre-Yves Ancel, Valérie Benhammou, Anass Ritmi, Laetitia Marchand, Cecile Zaros, Elodie Lordmi, Adriana Candea, Sophie de Visme, Thierry Simeon, Xavier Thierry, Bertrand Geay, Marie-Noelle Dufourg, Karen Milcent (Epipage2 and Elfe child cohorts)

Clovis Lusivika-Nzinga, Gregory Pannetier, Nathanael Lapidus, Frédéric Chau, Isabelle Goderel, Céline Dorival, Jérôme Nicol, Fabrice Carrat (IPLESP – methodology and coordinating data center)

Cindy Lai, Hélène Esperou, Sandrine Couffin-Cadiergues (Inserm)

Jean-Marie Gagliolo (Institut de Santé Publique)

Hélène Blanché, Jean-Marc Sébaoun, Jean-Christophe Beaudoin, Laetitia Gressin, Valérie Morel, Ouissam Ouili, Jean-François Deleuze (Fondation Jean-Dausset, CEPH-Biobank)

Stéphane Priet, Paola Mariela Saba Villarroel, Toscane Fourié, Souand Mohamed Ali, Abdenour Amroun, Morgan Seston, Nazli Ayhan, Boris Pastorino, Xavier de Lamballerie (Unité des Virus Émergents)
